# CT-based Osteoporosis Classification and Bone-Muscle Interaction Mapping Using Multiple Interpretable Machine Learning Models with the *BMINet* Framework

**DOI:** 10.1101/2025.02.12.25321163

**Authors:** Jingran Wang, Ziyan Hao, Liyun Lin, Jiachen Liu, Jiarong Wang, Zaixiang Tang, Dechun Geng, Caifang Ni, Huilin Yang, Kun Li, Jun Du

**Author notes:** **Correspondences: Jun Du**, Department of Orthopedic Magnetic Resonance Chamber, The First Affiliated Hospital of Soochow University, No. 899 Pinghai Road, Suzhou, 215000, China., **Kun Li**, Department of Orthopedic Surgery, The First Affiliated Hospital of Soochow University, No. 899 Pinghai Road, Suzhou, 215000, China., Biopharmagen Corp., Fangzhou Suzhou, No. 88 Dongchang Road, Suzhou 215127, P. R. China., **Huilin Yang**, Department of Orthopedic Surgery, The First Affiliated Hospital of Soochow University, No. 899 Pinghai Road, Suzhou, 215000, China., Biopharmagen Corp., Fangzhou Suzhou, No. 88 Dongchang Road, Suzhou 215127, P. R. China.

## Abstract

**Background:** Osteoporosis progresses through stages characterized by declining bone mineral density, vertebral deterioration, and muscle atrophy, with bone-muscle interactions driving synergistic degeneration.

**Methods:** This study retrospectively collected data from 444 patients aged 50 and older, who underwent DXA, CT, and MRI scans at the First Affiliated Hospital of Soochow University. CT values were measured for 6 vertebrae (L1-S1) and 30 adjacent muscle groups (psoas major, erector spinae, quadratus lumborum) to assess vertebral and muscle density. After analyzing changes in CT values across osteoporosis stages development to capture vertebrae and muscles degeneration pattern, we use multiple interpretable machine learning models to construct classification model and construct bone-muscle interaction network.

**Results:** This study found that osteoporosis progresses with age, with faster degeneration in females. Early stages show significant bone degradation, especially in L5 and S1 vertebrae, while later stages highlight muscle atrophy. Machine learning models, enhanced by Recursive Feature Elimination (RFE), effectively predicted disease progression (with Normal vs. Osteopenia 0.788, Normal vs. Osteoporosis 0.909, Normal vs. Osteoporotic fracture 0.942, Osteopenia vs. Osteoporosis 0.708, Osteopenia vs. Osteoporotic fracture 0.820 and Osteoporosis vs. Osteoporotic fracture 0.770). The Combined bone muscle interaction network reveals that vertebrae dominate early interactions, shifting to the muscle-clustered module in advanced stages, reflecting the complex degeneration of both bone and muscle.

**Conclusion:** This study develops classification models and analyze bone-muscle interactions in osteoporosis, uncovering synergistic degradation patterns across disease stages. The innovative BMINet toolkit offers an efficient, interpretable framework for personalized analysis, advancing precision medicine and integrated care for osteoporosis patients.

## Introduction

Osteoporosis and its most severe complication—osteoporotic fractures—have emerged as significant global public health challenges. This issue has become increasingly pressing with the rapid aging of populations worldwide.[1] Global statistics indicate that approximately one-third of women and one-fifth of men over the age of 50 will experience an osteoporotic fracture during their lifetime. These fractures often lead to long-term disability and loss of independence, significantly diminishing patients’ quality of life.[2, 3] At the same time, the high costs associated with acute fracture treatment, long-term care, and rehabilitation impose substantial financial burdens on patients’ families and healthcare systems.[4] Consequently, early diagnosis and effective intervention strategies to reduce the incidence of osteoporotic fractures have become critical issues in medical research and clinical practice.

The core pathological features of osteoporosis include a reduction in bone mineral density (BMD) and the degradation of bone microarchitecture. These deteriorations weaken the mechanical strength of bones, making them more susceptible to fractures.[5] While BMD decline is an important indicator for predicting fracture risk, an increasing number of studies have highlighted the critical role of complex physical and biological interactions between bone and muscle in the onset and progression of the disease.[6–9] In recent years, researchers have increasingly recognized that muscle atrophy, fat infiltration, and the concurrent degeneration of bone and muscle become particularly pronounced in the later stages of osteoporosis.[10] Therefore, relying solely on BMD for risk assessment has limitations, as overlooking changes in the muscular system may lead to insufficient risk prediction.

Although existing studies have revealed a close association between bone and muscle, current clinical practice still lacks efficient and systematic tools to analyze bone-muscle interactions[11]. This limitation constrains our comprehensive understanding of osteoporosis and leaves significant gaps in fracture risk assessment. Furthermore, existing research predominantly focuses on single BMD measurements, failing to fully capture the complex relationship between bone and muscle.[12] While some studies have begun using machine learning models to analyze osteoporosis data, effectively integrating multidimensional features of bone and muscle into a unified analytical framework for interaction analysis remains a pressing challenge.[13] Therefore, developing a systematic analytical method that can comprehensively integrate bone and muscle characteristics is essential for enhancing risk assessment, informing early intervention strategies, and advancing our understanding of the mechanisms behind bone-muscle co-degeneration.

CT values (Hounsfield Units, HU) are crucial indicators for assessing tissue density and X-ray absorption, widely used to evaluate bone and muscle health.[14, 15] Higher CT values typically correspond to denser tissues, such as bone, while lower CT values are associated with less dense tissues, such as fat or muscle.[16] A decline in CT values in bone suggests a reduction in bone mass or the onset of osteoporosis, while in muscle, lower CT values generally reflect muscle atrophy or quality loss.[17]

Our study systematically analyzed changes in bone and muscle CT values across different stages of osteoporosis and developed a classification model for osteoporosis stages.[18] We used interpretable machine learning to analyze bone-muscle co-degeneration and identify key nodes and interaction patterns in disease progression.[19] Furthermore, to support large-scale data analysis and the construction and analysis of bone-muscle interaction networks, we developed an innovative Python toolkit—BMINet. BMINet integrates various advanced machine learning algorithms, including RandomForest, XGBoost, LightGBM and CatBoost, and offers a comprehensive suite for data preprocessing, feature selection, model building and interpretation, as well as interaction network and pattern recognition. This tool not only enables an in-depth analysis of the complex interactions within the bone and muscle systems but also provides clinicians with a novel means of decision support.

In summary, our study not only advances the theoretical foundations of osteoporosis diagnosis and treatment but also introduces practical tools that can transform clinical practice. By fostering a more comprehensive understanding of the interplay between bone and muscle health, our research underscores the importance of a holistic approach to musculoskeletal disorders. This comprehensive strategy has the potential to lead to more integrated and efficient healthcare solutions, ultimately enhancing patient outcomes and reducing the overall burden of osteoporosis on individuals and healthcare systems alike.

## Methods

### Patients cohort

We retrospectively collected data from patients who underwent dual-energy X-ray absorptiometry (DXA), computed tomography (CT), and magnetic resonance imaging (MRI) at the First Affiliated Hospital of Soochow University from January 2016 to December 2022. MRI was used to confirm the presence of recent fractures. The inclusion criteria were as follows: patients aged 50 years and older who underwent the specified imaging exams on the spine. Exclusion criteria included incomplete patient records, lack of required imaging or diagnostic exams, presence of other diseases affecting bone density, or prior treatment for osteoporosis. The initial dataset consisted of 1,397 patients, of whom 656 were excluded due to a history of osteoporosis treatment. Upon further verification, 111 patients were excluded because of duplicate names or data errors. After a detailed review of imaging results, an additional 186 patients were excluded for reasons such as absence of lumbar spine CT scans (e.g., only cervical or thoracic CT was available), primary pathological fractures, or unreliable data measurement due to image artifacts (e.g., motion or foreign body artifacts). The final cohort included 444 patients, comprising 315 women (71%) and 129 men (29%). This study was approved by the institutional reviewboard of the First Affiliated Hospital of Soochow University (#2024-612), and individual informed consent for retrospective data collection was waived.

### Data collection and processing

All patients underwent scanning with a GE 256-slice spiral CT scanner (Revolution, GE Healthcare, USA) using the following parameters: tube voltage of 120 kVp, tube current of 499 mA, field of view of 500 mm, matrix size of 512×512, and a slice thickness of 3 mm, with images reconstructed using a soft tissue algorithm. DXA measurements were performed with a dual-energy X-ray absorptiometer (Hologic, USA), with patients lying supine on the examination table. Lumbar spine measurements were taken in the anteroposterior position. The scanning parameters for the lumbar BMD assessment included a tube current of 2.5–3.0 mA and tube voltage of 100–140 kV, with a scan width and length of 11.4 cm and 20.4 cm, respectively. BMD values (g/cm²) were calculated in the anteroposterior view, and the corresponding T-scores were determined according to diagnostic standards: T-score ≥ −1.0 SD indicated normal bone mass, −1.0 to −2.5 SD indicated low bone mass, < −2.5 SD indicated osteoporosis, and ≤ −2.5 SD with at least one fragility fracture indicated severe osteoporosis.

In CT image analysis, We measured CT values for six vertebrae (L1-S1) and the adjacent symmetrical muscle groups (Fig 1). At each disc level, four major muscle pairs—the Transversospinali, Lumbar spinalis, Lumbar longissimus and Psoas major—were measured bilaterally.[20] To facilitate measurement, the Transversospinalis and Lumbar spinalis muscles were combined, with the numbering rules defined as follows: Transversospinalis and Lumbar spinalis are numbered as 1 and 4, Lumbar longissimus muscle as 2 and 5, and Psoas major muscle as 3 and 6. The two numbers indicate spatially symmetrical muscles, and the naming convention specifies their location, such as L1-L2_1, which represents muscle number 1 situated between the L1 and L2 vertebrae layer.

**Fig. 1:**
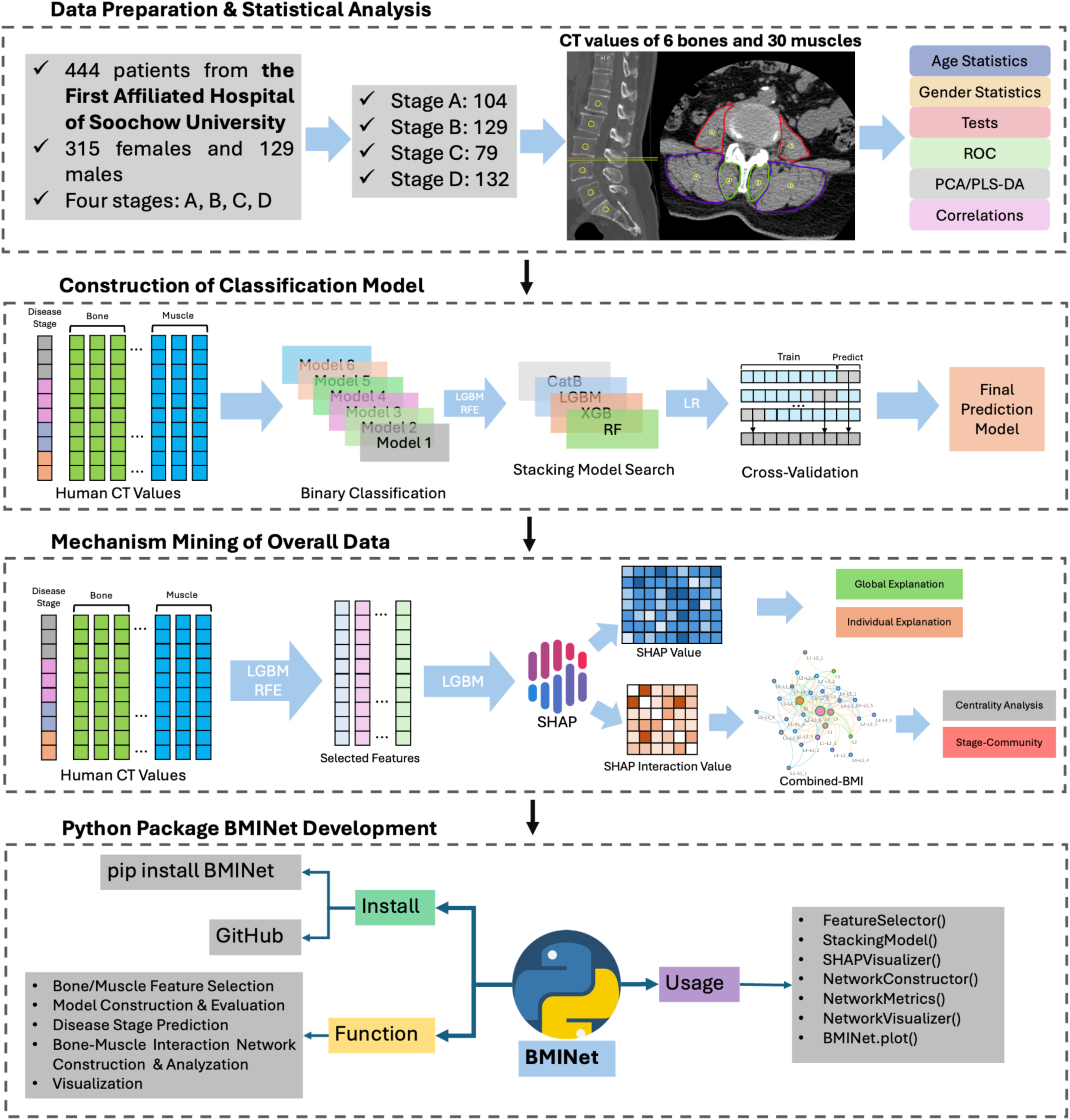
Flow chart of the overall study design. Our study includes data preparation, construction of classification models, construction of codegenerate interactive networks, and Python package development.

Regions of interest (ROIs) were selected according to strict criteria: for vertebrae, ROIs were placed within the central area of each vertebral body, avoiding the cortical bone region to ensure accurate bone density readings. For muscle CT value measurements, ROIs were placed in the center of each muscle, avoiding any interference from bone signals that could introduce partial volume effects. Each ROI had an approximate area of 150 mm², and one ROI was positioned in each of the three central slices at each disc level. For each vertebra and each muscle, the average of three ROI measurements was calculated as the final CT value, ensuring data accuracy and consistency.

Overall, we measured CT values for six vertebrae and 30 muscles, yielding CT values for 36 bone-muscle features (see Fig 1 for the measurement method, and Supplementary Table 1 for the original CT values). These data were subsequently analyzed to assess changes in vertebral and muscle CT values across different osteoporosis stages and to compare variations among groups.

### Statistical analysis

Data analysis was conducted using Python version 3.10 (https://www.python.org) and R version (https://www.r-project.org). Visualizations were performed using the matplotlib package (version 3.7.2) and seaborn package (version 0.12.2) in Python, as well as the ggplot2 package (version 3.5.1) and corrplot package (version 0.92) in R.[21–23] Statistical tests were conducted across different groups to investigate trends in bone and muscle CT value characteristics. For normally distributed data, two-tailed t-tests were applied, for data that did not meet normal distribution criteria or had small sample sizes (fewer than 30 non-missing values), Wilcoxon rank-sum tests were used for nonparametric analysis. Model performance was evaluated using receiver operating characteristic (ROC) curves, with primary metrics including True Positive Rate (TPR), False Positive Rate (FPR), and Area Under the Curve (AUC). The formulas for these metrics are as follows:

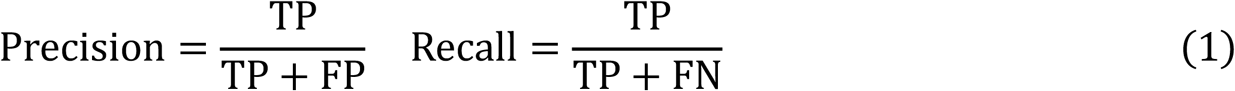

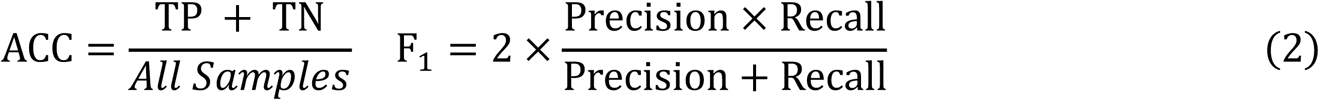

In these formulas, TP represents True Positives, TN represents True Negatives, FP represents False Positives, and FN represents False Negatives. AUC (Area Under the Curve) reflects the model’s overall performance in binary classification tasks. For each individual feature, we plotted ROC curves and compared them across disease groups. In the single-feature ROC analysis, we excluded NA values and compared AUC values to determine the relative importance of features in distinguishing different disease states. In the dimensionality reduction and correlation analysis, NA values were filled using group-wise mean imputation.We applied two dimensionality reduction techniques—Principal Component Analysis (PCA) and Partial Least Squares Discriminant Analysis (PLS-DA)—to explore the distribution differences of CT value data across different groups. Using these methods, we observed distributional differences among disease groups in the reduced dimensions, revealing potential patterns in the disease progression process.[24] Additionally, we used Spearman’s correlation analysis to construct a correlation matrix between vertebrae and muscles, followed by hierarchical clustering of features using the maximum distance method. This approach helped us explore the relationships between features and preliminarily assess the degeneration trends between bone and muscle, identifying bone-muscle groups that share similar degeneration patterns.[25]

### Feature Selection & Model Construction

Through data measurement, we obtained CT value for 6 vertebrae and 30 muscles, along with patient age and gender information. In the analysis involving explainable machine learning methods, considering that missing values (NA) in CT measurements, and to facilitate future sample studies, we used machine learning methods capable of automatically handling missing data without imputing the NA values in the original data. To efficiently find the optimal feature combination and effectively handle missing data, we first used Light Gradient Boosting Machine (LightGBM) as the core algorithm for feature selection. LightGBM is an efficient gradient boosting algorithm that improves model prediction performance by building a series of decision trees. We applied Recursive Feature Elimination (RFE), which works by gradually eliminating features with lower importance to optimize the feature set. During training, RFE evaluates the importance of each feature in each iteration, progressively discarding those with less contribution to the model, ultimately identifying the best feature combination. Model performance was evaluated using 5-fold cross-validation, and the average accuracy of the model was computed for each binary classification task.

In addition to the LightGBM model, we also incorporated three other tree-based algorithms: Random Forest, eXtreme Gradient Boosting (XGBoost), and Categorical Boosting (CatBoost). We combined these four algorithms and explored various learning rates and hyperparameter combinations. Additionally, we used logistic regression as a meta-model to construct a stacking classifier. The stacking model integrates the predictions of multiple base models by training a meta-model on their outputs, thereby improving overall classification performance through generalization. The stacking model’s performance was evaluated using 5-fold cross-validation, where we calculated the maximum AUC for each binary classification task. By exploring different combinations of base models, we automatically compared the performance of model combinations and selected the best combination for the final model. Model training and evaluation were performed using Python’s scikit-learn (version 1.3.0), XGBoost (version 2.1.1), LightGBM (version 4.5.0), and CatBoost (version 1.2.5).[26–29]

### SHAP based model explanation

Machine learning models are often used as a “black box,” but through explainable machine learning, we can open this black box and uncover the underlying mechanisms.[30] To address this issue, we applied the SHAP (SHapley Additive exPlanations) method to interpret the machine learning model and identify the key features and medical mechanisms involved in the classification predictions for osteoporosis and related diseases.[31] SHAP is a powerful explanation method in machine learning that provides both global feature importance rankings and explanations for individual predictions. This method breaks down each sample’s prediction into contributions from individual features, clearly showing which features had the greatest influence on the prediction for that sample.[32]

For each binary classification task at different disease stages, we used the optimal feature combinations selected during the Feature Selection step. We then split these features into an 80% training set and a 20% validation set, trained a LightGBM model, and input the trained model into SHAP for both global and individual explanations of the entire dataset. Through SHAP values, we were able to precisely identify which vertebrae and muscle CT values are crucial for distinguishing disease progression across different stages, including normal bone density, reduced bone mass, osteoporosis, and osteoporotic fractures. Furthermore, we used SHAP dependency plots to display the interactions between vertebrae and muscle features in the classification models for different disease stages, revealing potential interactions between two features.[33]

### Bone-Muscle interaction detection

To further explore and quantify the complex interactions between bones and muscles, we introduced the concept of SHAP interaction values to reveal the interaction effects between pairs of features in the model’s predictions.[34] SHAP interaction values explain the role of feature interaction in two steps: first, evaluating each feature’s contribution to the prediction independently, and then calculating the interaction effect between it and other features, which refers to how the joint presence of both features influences the prediction outcome.[35] The SHAP Interaction Value can be calculated using the following formula based on game theory:

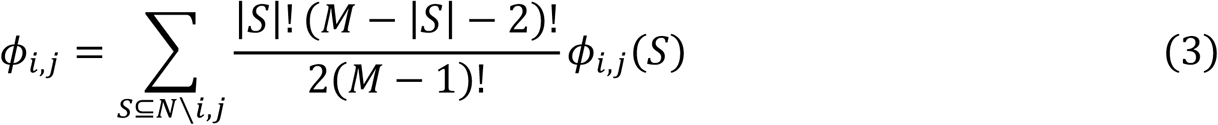

where *ϕ_i,j_* represents the interaction effect between feature *i* and feature *j* and *S* represents all subsets without feature *i* and feature *j* and *ϕ_i,j_*(*S*) is the output difference of the model after adding features i and j versus adding only i or j can be calculated as *ϕ_i,j_*(*S*) = *f*(*S* ∪ {*i*, *j*}) − *f*(*S* ∪ {*i*}) − *f*(*S* ∪ {*j*}) + *f*(*S*). Where *f*(*S*) is the model output under subset *S*. Using the LightGBM model, we calculated the SHAP interaction values for the vertebrae and muscle features in the classification models at different disease stages. This allowed us to identify which features have significant interaction effects in the progression of disease stages.[36] By analyzing these interaction effects, we were able to uncover the collaborative action patterns of vertebrae and muscle features at different stages of disease development.

### Interation network construction and analysis

To reveal the interactions pattern between bones and muscles in the progression of osteoporosis, we constructed an interaction network based on SHAP interaction values. Firstly, the nodes of the network were composed of significant bone and muscle features identified by the LightGBM model. Then, we calculated the SHAP interaction values between each pair of features. In this study, a cutoff value of 1.5 was set, and interactions with a value greater than this threshold were considered significant and incorporated as edges into the interaction network. We then integrated the sub-networks constructed for each stage comparison into a “Combined Bone-Muscle Interaction Network” (Combined-BMI) to evaluate the global representation of bone-muscle interactions during disease progression. We then calculated three centrality parameters for each node in the network: Betweenness, Closeness and Degree. These parameters are used to measure the importance of nodes in the communication within the network. Betweenness Centrality reflects the frequency with which a node appears on the shortest paths, representing its importance as an information hub in the network.[25] The formula is:

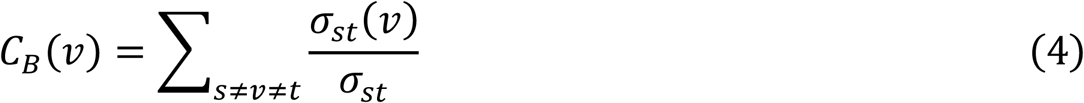

Where *σ_st_* is the number of shortest paths from node s to node t, and *σ_st_*(*v*) is the number of shortest paths passing through node v. Closeness Centrality measures the average shortest path length from a node to all other nodes in the network, reflecting the communication efficiency of the node. A larger Closeness value indicates that communication from this node to others is relatively easy.[37] The formula for Closeness Centrality is:

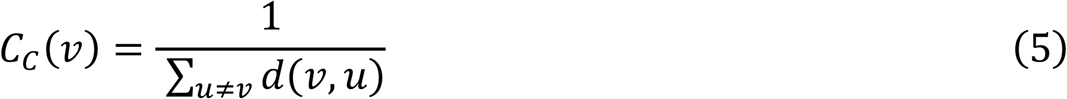

where *d*(*v*, *u*) is the shortest path distance from node v to node u. Degree represents the number of edges connected to a node. The higher the degree, the more connections the node has, and the more important it is in the network. To further reveal the interaction modules between bone and muscle CT value features in the progression of osteoporosis, we used the Louvain community detection algorithm. This algorithm optimizes community division by maximizing modularity, dividing the nodes in the interaction network into multiple tightly connected subsets.[38] The calculation formula for modularity Q is:

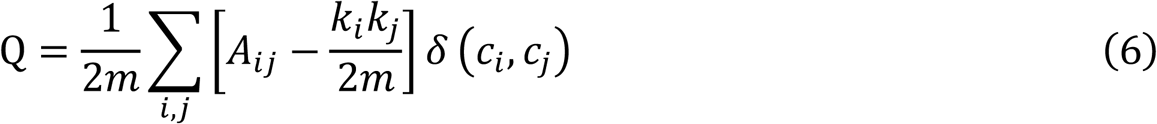

where *A_ij_* is the adjacency matrix, *k_i_*, *k_j_* are the degrees of nodes i and j, mmm is the total number of edges in the network, and *δ*(*c_i_*, *c_j_*) is an indicator function that takes the value 1 if nodes i and j belong to the same community, and 0 otherwise. The construction and analysis of the interaction network were completed using the networkx package (version 2.8.8) in Python, and the network visualization was presented using Cytoscape software (version 3.10.2) to display the complex bone-muscle interaction structure.[39, 40]

## Results

### Patient characteristics

In this study, based on the bone mineral density (BMD) diagnostic results and whether osteoporotic fractures occurred, we divided the 444 patients’ CT images into four stages: Stage A (Normal group, n=104), Stage B (Osteopenia patients, n=129), Stage C (Osteoporosis patients, n=79), and Stage D (Osteoporotic fracture patients, n=132) (Fig 2a). The gender distribution showed 315 female patients, accounting for 71%, and 129 male patients, accounting for 29% (Fig 2b). Age distribution analysis using kernel density estimation revealed that the participants’ ages were concentrated between 50 and 85 years. With the progression of the disease, from osteopenia to osteoporosis and osteoporotic fractures, the peak age gradually shifted to an older range. Specifically, the peak age for Stage B was 63.17 years (95% CI: 52.20-81.80), for Stage C was 68.21 years (95% CI: 50.95-84.00), and for Stage D, the peak age reached 73.02 years (95% CI: 53.27-88.45) (Fig 2c). These age trends suggest that the progression from osteopenia to osteoporosis, and then to osteoporotic fractures, takes approximately 5 years, and the severity of the disease gradually increases with age.

**Fig. 2:**
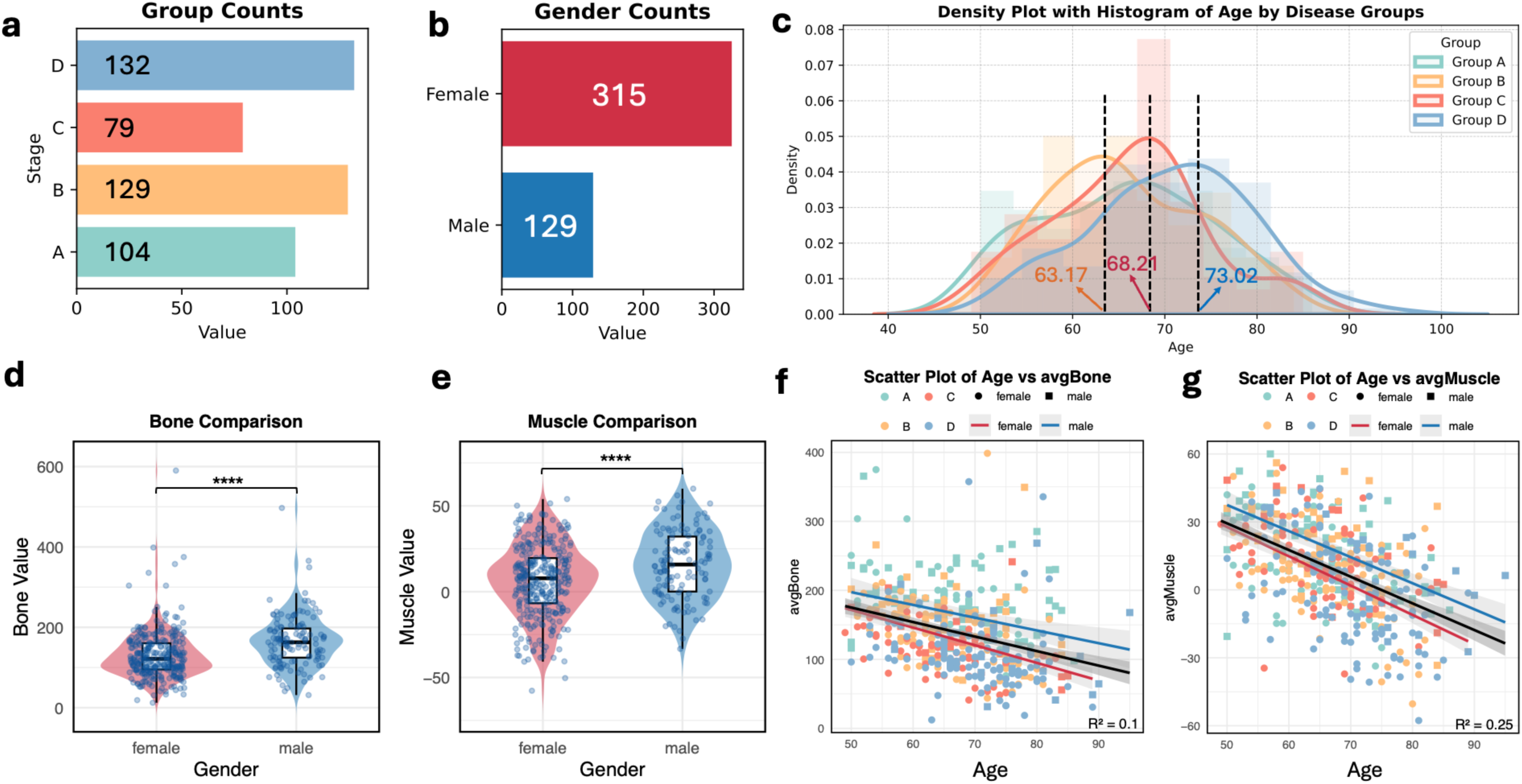
Data Distribution and Patient Characteristics. (a) Sample grouping and distribution of sample sizes. (b) Gender distribution. (c) Kernel density estimation of age distribution in each group. (d) Female average vertebral CT values are significantly lower than males (p ≤ 0.0001). (e) Female average muscle CT values are significantly lower than males (p ≤ 0.0001), indicating reduced bone mass and muscle mass. (f) Trend of vertebral CT average values across age for each group, with a faster decline in females compared to males. (g) Trend of muscle CT average values across age for each group, with a more significant decline in females. *: 0.01 < p ≤ 0.05, **: 0.001 < p ≤ 0.01, ***: 0.0001 < p ≤ 0.001, ****: p ≤ 0.0001.

We further analyzed the distribution of average vertebral and muscle CT values in different genders and explored the impact of age on these features. The results indicated that women had lower vertebral and muscle CT values compared to men, with a statistically significant difference (p < 0.0001). (Fig 2d, e). Additionally, as age increased, both vertebral and muscle CT values showed a significant decreasing trend, with a faster rate of decline in female patients compared to male patients (Fig 2f, g).

### Statistical analysis of different disease stages

To assess the trend of changes in vertebral and muscle CT values at different stages of osteoporosis (from normal to osteoporotic fractures), we performed pairwise comparisons of 36 individual CT values across the four stages. We selected vertebral and muscle features with significant differences (p-value < 0.05) between stages (Supplementary Fig 1). Detailed statistical methods and the significance test results for each feature are presented in the supplementary materials (Supplementary Table 2). The results showed that the CT values of the L5 and S1 vertebrae exhibited significant changes throughout the disease progression, particularly during the early stage from normal to osteopenia. The significant changes in vertebral CT values mainly occur during the early stages of the disease, particularly at L3, L4 and L5, while changes in the later stages are relatively mild. From normal to osteopenia and from osteopenia to osteoporosis, the changes in muscle CT values were not significant. As the disease progressed from osteoporosis to osteoporotic fractures, the decline in all muscle CT values became significant, indicating that the degernations in muscles are more pronounced in the later stages of the disease.

### Performance of Vertebral and Muscle CT Values at Different Stages

To comprehensively evaluate the classification diagnostic ability of the 36 features of the vertebrae and muscles in different disease stages, we quantified each feature using the Receiver Operating Characteristic (ROC) curve and Area Under the Curve (AUC) after removing NA values (Fig 3a). The results showed that some vertebral features exhibited high classification performance, particularly the L5 vertebra which achieved an AUC of 0.921 in distinguishing between Stage A and Stage D. Additionally, L4 also demonstrated high AUC values (>0.7) in differentiating between Stages A and Stage B, and Stage A and Stage C, suggesting that L5 and L4 are key biomarkers of disease progression (Supplymentary Table 3). When the disease progressed to the later stages (i.e., comparison between Stages C and Stage D), the classification performance using only vertebral CT values declined. Certain muscle features (e.g., L1-L2_6) showed better performance with an AUC value of 0.743.

**Fig. 3:**
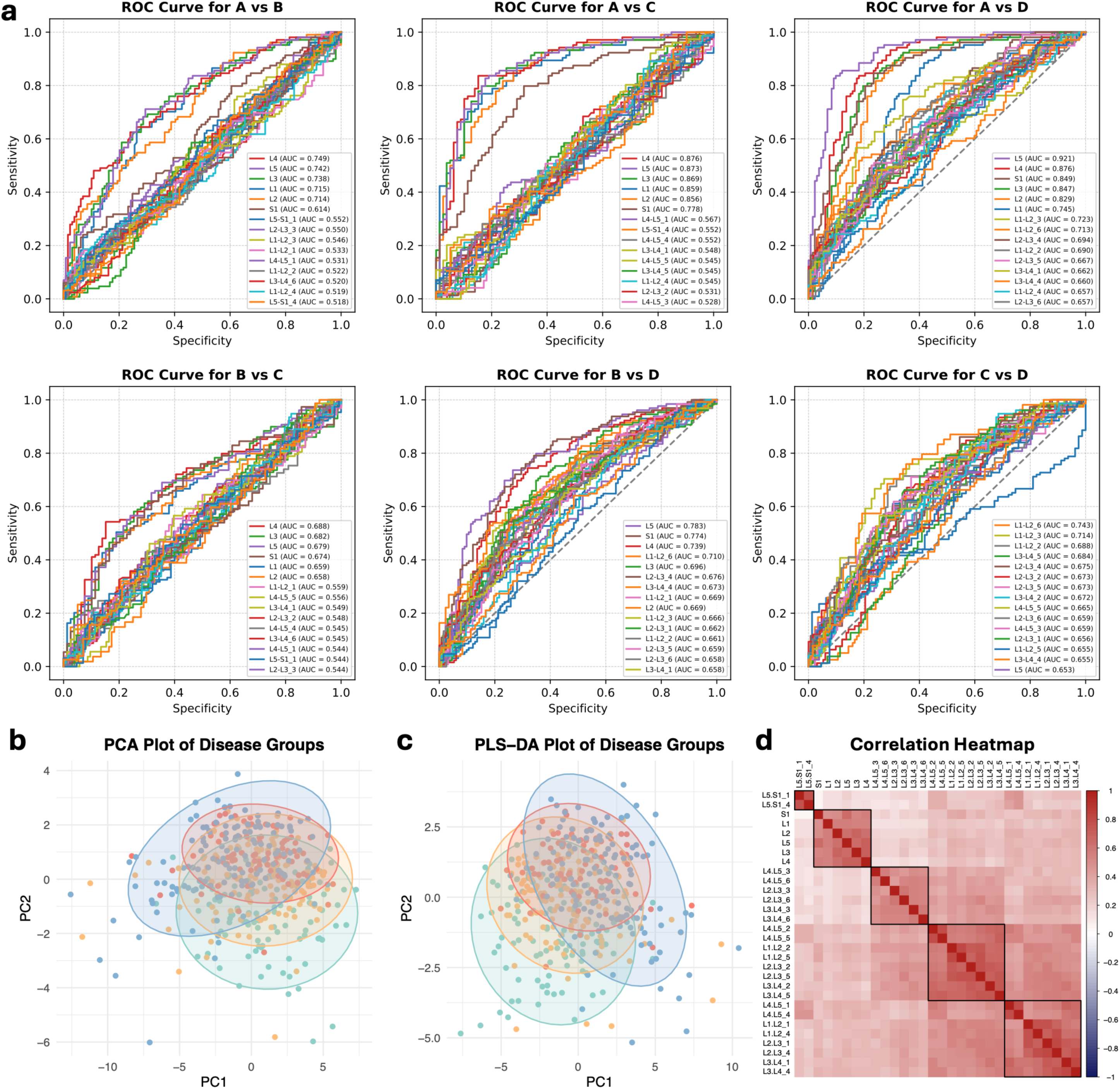
Classification performance, dimensionality reduction, and correlation analysis of CT values. (a) ROC curves quantifying classification performance across disease groups. (b) PCA reveals distribution trends, with increasing separation between C and D groups as disease progresses. (c) PLS-DA further confirms feature differentiation across stages. (d) Correlation heatmap shows significant relationships between vertebra and muscle features, highlighting their synergistic degeneration.

### Severe disease stages show more significant CT features

To further analyze the distribution patterns of the 36 vertebral and muscle CT features across different disease stages, we applied Principal Component Analysis (PCA) and Partial Least Squares Discriminant Analysis (PLS-DA) for dimensionality reduction (Fig 3b, c). The results showed substantial overlap between Stage A, Stage B and Stage C along PC1, indicating limited feature changes in the early disease stages. As disease stage devveloped, a gradual separation emerged between Stage C and Stage D PC1, suggesting a divergence in CT features related to bone-muscle degeneration. PLS-DA as a supervised method, more clearly revealed the feature separation between Stage C and Stage D along both PC1 and PC2, indicating more prominent changes in CT features as the disease progressed to later stage (fracture phase).

### Vertebraes and symmetric muscles exhibit similar degenerative characteristics

We further explored the intrinsic relationships between different CT features using Spearman correlation analysis to construct correlation heatmap (Fig 3d). The highly correlated features show the synergistic degeneration of bone and muscle in Osteoporosis progression. We identified five significant clusters: one consisting of all vertebrae, one containing muscles 2 and 5, one consisting of muscles 3 and 6, one with all muscles 1 and 4 except for L5_S1_1 and L5_S1_4, and a final one containing only L5_S1_1 and L5_S1_4. This suggests that during the overall vertebral-muscle degeneration process, the degeneration of vertebrae exhibits similar characteristics, while the degeneration of spatially symmetric muscles also shares some similarities. Notably, the degeneration characteristics of the L5-S1_1 and L5-S1_4 muscles are relatively unique compared to other muscles 1 and 4, as they exhibit a lower degree of degeneration throughout the development of osteoporosis.

### Indicators Detection and Classification Performance Evaluation

In disease stage classification, the performance of individual vertebra and muscle CT values is limited by low AUC and insufficient predictive accuracy, as well as the presence of NA values in some features. Additionally, relying solely on single features limits the model’s overall performance due to incomplete feature utilization. We applied Recursive Feature Elimination (RFE) combined with multiple machine learning models for comprehensive feature selection and performance optimization (Fig 4).

**Fig. 4:**
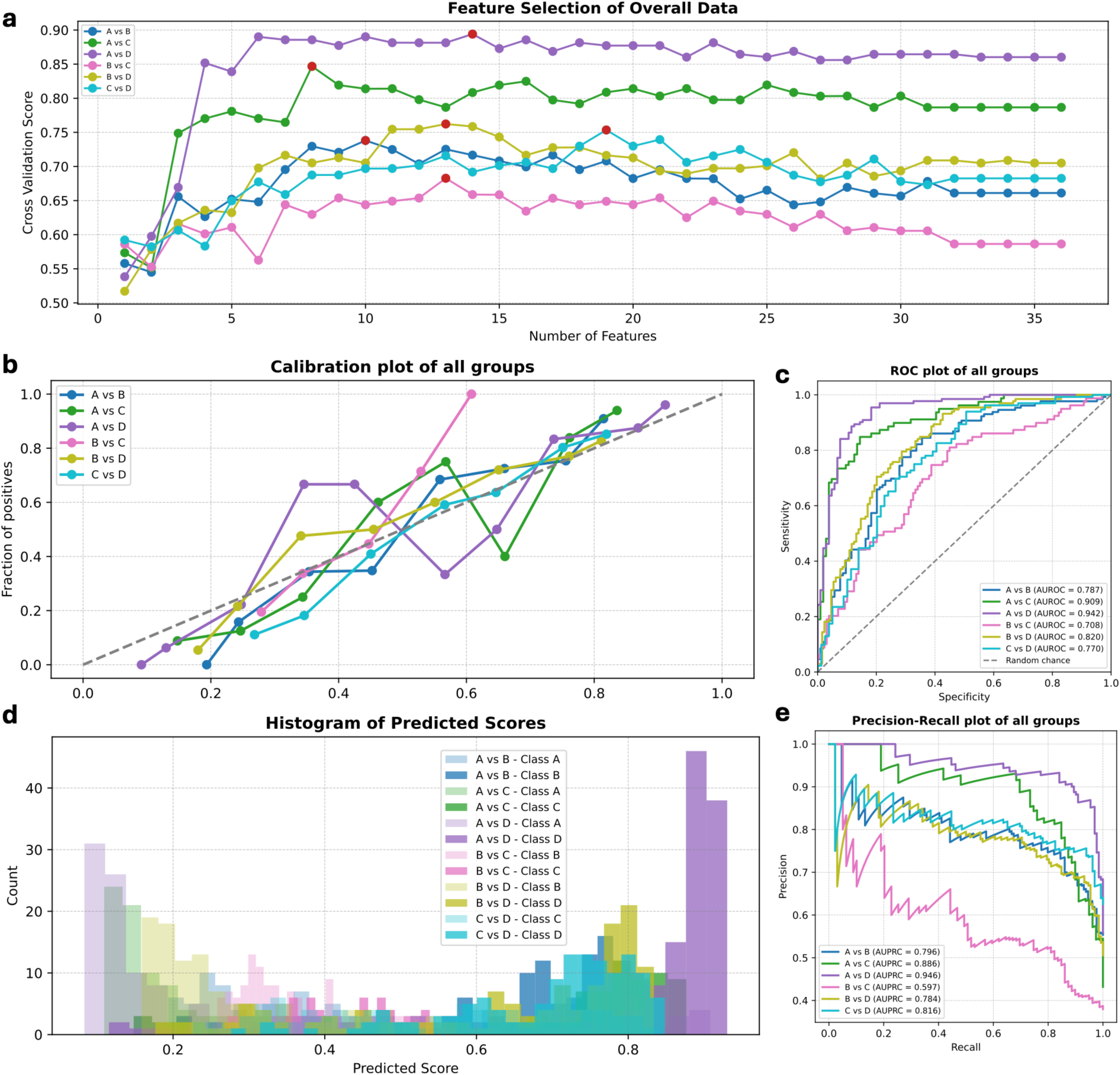
Machine learning models construction. (a) Using RFE to identify indicators, with the best feature combinations selected via cross-validation scores. (b) Calibration plot of the integrated indicators obtained from the selected features. (c) ROC curve of the integrated indicators. (d) Distribution histogram of the integrated indicators. (e) Precision-Recall curve of the integrated indicators.

By controlling for confounding factors such as gender and age, we focused on the CT value changes of vertebrae and muscles. To efficiently identify the best feature combinations from datasets with NA values, we compared RFE methods based on LightGBM and XGBoost, finding that LightGBM outperformed XGBoost (Supplementary Table 4). Using LightGBM, we progressively eliminated low-importance features and evaluated cross-validation scores between disease stages, identifying the optimal feature set for vertebrae and muscle combinations (Fig 4a). Using this feature set, we applied a Stacking model for all disease stages classification (Table 1). Calibration curves (Fig 4b) showed that the calibration for A vs. D was better than other groups. The ROC curves (Fig 4c) displayed the performance of classification models for each stage comparasion, with A vs. D and A vs. C showing the best performance (AUROCs of 0.942 and 0.909, respectively). Conversely, the B vs. C classification had a lower AUC (0.708), indicating that differentiating between bone loss and osteoporosis is more challenging. The histogram of predicted scores (Fig 4d) revealed significant differences in the distribution of predicted probabilities. Additionally, Precision-Recall curves (Fig 4e) assessed the performance of different models on imbalanced data, confirming the model’s superiority in complex classification tasks. Furthermore, integrating multiple features (without excluding NA values) yielded higher AUC than the best single feature with NA values excluded, indicating that combining multiple parameters enhances classification performance.

**Table 1:** Best Models and Performance Metrics for Comparisons Among Different Groups. LGBM represents LightGBM (Light Gradient Boost Machine), CatB represents CatBoost (Categorical Boosting), RF represents Random Forest, XGB represents XGBoost (eXtreme Gradient Boosting), AUROC represents area under ROC curve, AUPRC represents area under precision-recall plot and ACC represents accuracy.

| Group | Indicators | Model | AUROC | AUPRC | ACC | Precision | Recall | F1 |
| --- | --- | --- | --- | --- | --- | --- | --- | --- |
| A vs B | 10 | LGBM & CatB | 0.788 | 0.796 | 0.742 | 0.748 | 0.806 | 0.776 |
| A vs C | 8 | LGBM & RF | 0.909 | 0.886 | 0.842 | 0.821 | 0.810 | 0.815 |
| A vs D | 14 | LGBM & RF | 0.942 | 0.946 | 0.873 | 0.870 | 0.909 | 0.889 |
| B vs C | 13 | LGBM & RF | 0.708 | 0.597 | 0.668 | 0.727 | 0.203 | 0.317 |
| B vs D | 13 | LGBM & XGB & CatB | 0.820 | 0.784 | 0.747 | 0.754 | 0.742 | 0.748 |
| C vs D | 19 | RF & XGB & CatB | 0.770 | 0.816 | 0.744 | 0.747 | 0.894 | 0.814 |

### Indicators explanation indicates disease stages development mechanism

Given the predictive performance of the LightGBM model, we further applied SHAP (Shapley Additive Explanations) to deeply analyze the model’s predictions, revealing feature importance and variation patterns across disease stages. The SHAP heatmap of global model explanations showed that in comparisons between the normal and disease groups, vertebrae L4 and L5 exhibited high importance, where declines in their values drove the model to classify samples as disease states (Fig 5a-c). As the disease progressed from Stage B to Stage C, the importance of vertebrae S1 gradually increased and L5 remains its critical position (Fig 5d-e). Notably, in the late disease stages (Stage C to Stage D), the importance of vertebral features decreased, with the model relying more on muscle features like L1-L2_2 and L1-L2_3 for classification (Fig 5f). This shift suggests that vertebral changes are critical in the early stages, while muscle feature changes become more significant for late-stage disease classification.

**Fig. 5:**
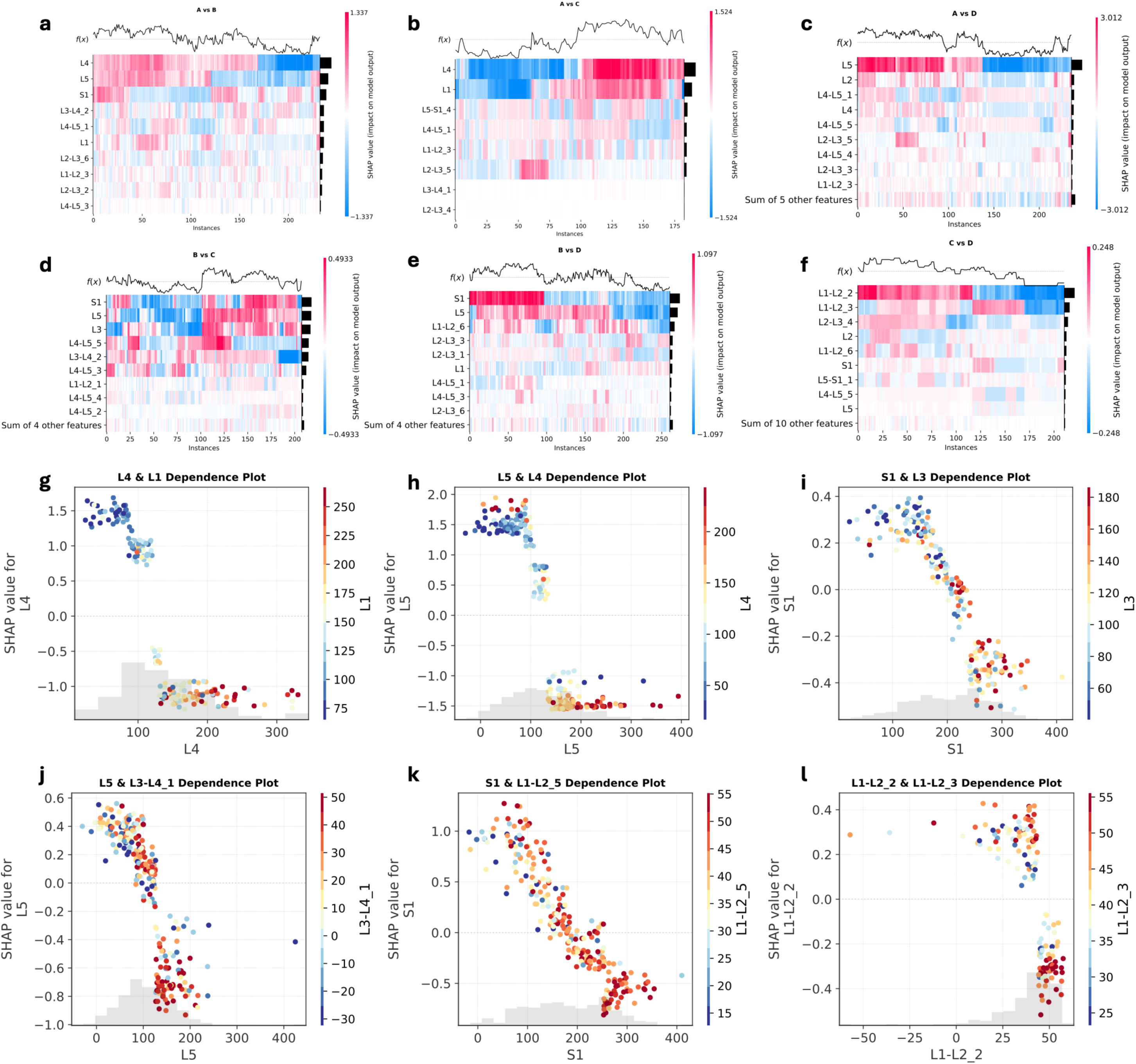
SHAP heatmaps and SHAP dependency plots. (a-f) SHAP value heatmaps for the LightGBM model, highlighting feature impact across groups. Important features are positioned higher, reflecting greater influence on predictions. SHAP values are color-coded by magnitude and direction. The function f(x) at the top represents the model’s predicted output, illustrating how features contribute to the prediction. (g-l) SHAP dependence plots illustrates the relationship between the SHAP values for feature (y-axis) and the values of itself (x-axis), while also showing the interaction effect of another feature through color coding, where warmer colors indicate its higher values.

Beyond single-feature importance, the model captured interactions among features. The SHAP interaction effect plots illustrate not only each feature’s SHAP value relative to its CT value but also use color coding to show interactions with another feature (Fig 5g-l). For instance, in the interaction dependency plot between L5 and L4 (Fig 5h), as L5 values decrease, the model increasingly predicts more severe disease stages, a trend that is further amplified when L4 values also decrease. This reveals that the model considers not only individual feature changes but also the complex relationships among multiple features.

### Bone-muscle interaction network reveals stage development pattern

To quantify the interaction strength between different features and identify the significant interaction effects between bone and muscle characteristics, we computed SHAP Interaction Values and constructed a global bone-muscle interaction network (Fig 6a). This network reveals the complex developmental mechanisms of osteoporosis and related diseases at different stages. Based on this network, we constructed disease-specific sub-networks with vertebral and muscle features as nodes and interactions as edges. Using the Louvain community detection algorithm, three major communities were identified (Figure 6b, c). By calculating the betweenness centrality, closeness centrality, and degree of each node (Fig 6d), we clarified the importance of each node in the network:

1. **Community 1**: Centered on L5, L4, and L1 vertebrae, primarily demonstrating bone-bone and bone-muscle interaction patterns.
2. **Community 2**: S1 vertebra serves as the core node, with bone-bone interactions decreasing and muscle-muscle interactions gradually increasing, with muscle features like L1-L2_2 serving as bridge nodes between Communities 2 and 3.
3. **Community 3**: Composed entirely of muscle interactions with all nodes are muscle, where nodes within this community show relatively high degrees but lower betweenness values.

**Fig. 6:**
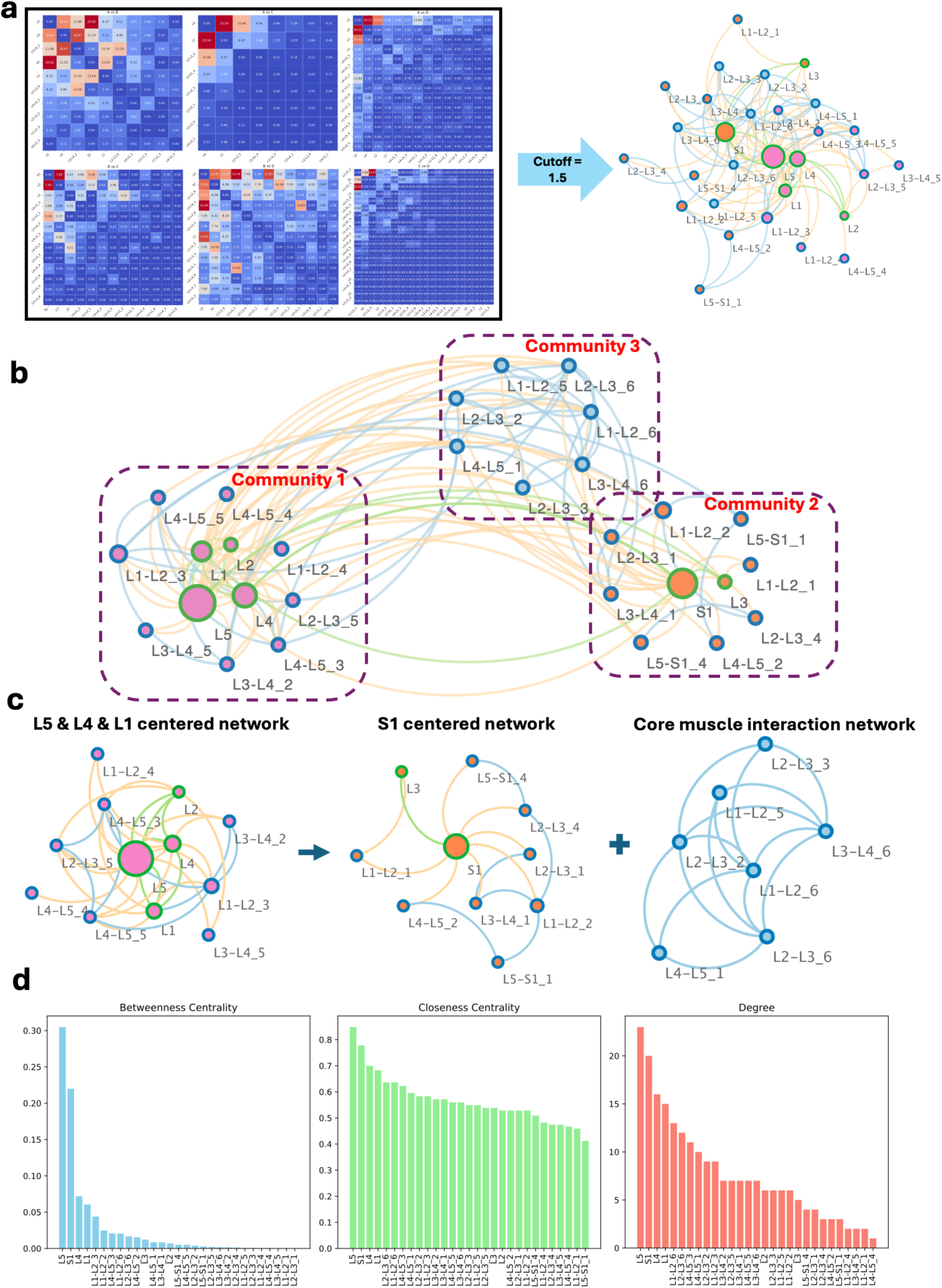
Construction and Analysis of the Combined-BMI Network. (a) Visualization of the Combined-BMI network, where three communities are detected using the Louvain community detection method. (b) The adjacency matrix of the network, with edges shown in blue and absence of edges in white. (c) The three identified communities: L5 & L4 & L1 centered network, S1 centered network, and core muscle interaction network. (d) Centrality measures of the nodes in the network, showing Betweenness Centrality, Closeness Centrality, and Degree values for each node.

The results indicate that L5 vertebra consistently ranks highest across all parameters, suggesting its dominant role in network communication, followed by S1 vertebra. Among muscle features, L1-L2_3, L2-L3_6, and L1-L2_6 rank highest in betweenness, closeness centrality, and degree, indicating their key roles in both local and global interactions. Overall, by constructing the Combined-BMI network, we have uncovered the collaborative degradation characteristics and patterns across osteoporosis stages develpoment. Specifically, we found that the communities identified in the Combined-BMI network are closely associated with the development of osteoporosis and its stages.

Community 1’s core nodes, including L5, L4, and L1 vertebrae, exhibit significant degradation throughout the stages of osteoporosis progression. These nodes are of high importance in distinguishing between the normal group and other disease stages, indicating that Community 1 represents the key interaction pattern in the transition from a healthy state to the degenerative phase of osteoporosis. Furthermore, the betweenness values of the nodes in Community 1 remain relatively high throughout disease progression, reinforcing the central role of these nodes in the disease development process. In Community 2, the interaction pattern centered around the S1 vertebra demonstrates its growing importance as osteoporosis progresses from Stage B to later stages. This indicates that the interactions in Community 2 are characteristic of the transition from Stage B to more severe stages of the disease. As the disease progresses to Stage C and Stage D, muscle characteristics become more critical for classification, surpassing the importance of the vertebrae. During this phase, Community 3, which is composed entirely of muscle interactions, plays a significant role. Notably, the L1-L2_2 muscle node in Community 3 becomes crucial for classification in Stages C and D, bridging Community 2 and Community 3 muscle-dominant interaction network. Meanwhile, the importance of the S1-centered interactions in Community 2 decreases.

It is noteworthy that the vertebrae and muscles, as interaction hubs, exhibit clear signs of degradation during the progression of osteoporosis. This phenomenon suggests that as the disease progresses, nodes that initially serve as key interaction hubs begin to degrade and lose their central role in signal transmission, leading to a decline in their functional significance. The degradation of the vertebrae and muscles is closely related to their changing roles in the disease progression, with their participation and importance shifting at different stages. In terms of network structure, this transformation is reflected in the transition of interaction communities: Initially, the communication relies on vertebrae as central nodes, forming a global vertebra-centered communication network with high betweenness values. As the disease progresses, the role of the vertebrae nodes weakens, and the network shifts towards a more localized interaction module centered around muscles, with higher degree nodes taking precedence. Additionally, significant vertebral nodes serve global communication functions in the transition from a normal state to osteoporosis, where vertebrae play a crucial role in network connectivity. As the disease progresses into osteoporosis and osteoporotic fractures, the muscle interactions begin to dominate communication in local regions, with muscles gather as cluster to play a critical coordinating role, especially during the transition from osteoporosis to osteoporotic fractures. These shifts are not only changes in interaction patterns but also reflects the physiological complexity of the disease. The degeneration and functional loss of vertebrae may prevent them from continuing to serve as central nodes in the later stages, while muscles gradually form local clusters and take over the key roles in communication and regulation.

### Usage of the BMINet Python Package

The BMINet Python package offers a range of core functions and features, enabling flexible usage and secondary development. Key functionalities include:

1. **Indicators Selection and Optimization**: BMINet integrates an automated feature selection function to ensure optimal model performance during training and prediction.
2. **Model Building and Prediction**: Users can choose from various machine learning models suitable for binary or multi-class classification tasks.
3. **Bone-Muscle Interaction Network Construction**: The built-in functions of BMINet enable fast construction of bone-muscle interaction networks, revealing complex relationships between bones and muscles during disease progression.
4. **Network Analysis**: To automated calculate network centrality parameters and detect communities.

## Discussion

This study analyzed CT values from 444 patients to explore osteoporosis progression and the co-degradation of bones and muscles. It found that the disease evolves over approximately five years, with age being a key factor and women experiencing faster degeneration than men. Machine learning models confirmed the combined predictive value of bone and muscle features, with RFE improving classification and SHAP providing model interpretability. Early stages highlighted vertebral CT value declines (L5, S1) as biomarkers, while later stages emphasized muscle atrophy’s role in fracture risk. A Combined-BMI network revealed a shift from vertebra-centered global interactions to muscle-dominated local clusters, reflecting the disease’s complexity and progression. This study systematically elucidates the co-degradation phenomenon of the skeletal and muscular systems, demonstrating that muscle atrophy and fat infiltration are significant predictors of fracture risk and act synergistically with bone density loss. Recent studies indicate that in osteoporosis, the L4 and L5 vertebrae experience greater stress during flexion and lateral bending,[41] while other research shows marked degeneration of the L4 and L5 vertebrae in osteopenia and osteoporosis.[42] In terms of muscles, studies have also established the correlation between muscle mass and osteoporosis. Consistent with these findings, our research emphasizes the importance of integrated management of bones and muscles in the prevention and treatment of osteoporotic fractures.[43]

The clinical translation potential of BMINet lies in its ability to bridge risk prediction and actionable interventions, offering a comprehensive tool for osteoporosis management. The significant decline in L5 vertebral CT values during the transition from normal to osteoporotic fractures provides a cost-effective screening indicator. BMINet enables automated risk stratification, making it highly applicable in primary healthcare facilities. Furthermore, the increased predictive weight of muscle features, such as L1-L2_3, in the fracture stage highlights the potential of targeted muscle-strengthening exercises to slow disease progression. Dynamic CT value monitoring, combined with rehabilitation efficacy assessment, could optimize resistance training protocols, offering a personalized approach to intervention. Additionally, the co-degeneration pattern may guide the development of novel dual-target therapeutic strategies. Preclinical studies have demonstrated that simultaneously targeting bone formation and muscle metabolism can yield synergistic effects,[7] and BMINet’s interaction network analysis provides a data-driven foundation for selecting such therapeutic targets. This framework not only enhances risk prediction but also bridges the gap between diagnostic insights and actionable interventions, paving the way for precision medicine in osteoporosis management. Furthermore, the BMINet framework is not limited to osteoporosis or CT-related research; it is designed to be adaptable for analyzing interaction patterns across various types of biological data.

This study also has several limitations. First, the sample predominantly consists of patients aged 50 to 85 years, with an uneven gender distribution—71% female—which may introduce bias in gender difference analysis and limit the generalizability of the results, particularly for younger populations. Second, some cases contain missing values (NA values). Although we addressed this issue through data imputation and partial processing, incomplete data might still influence the model outcomes. Moreover, the interaction networks lack experimental validation. While the interaction patterns were inferred from model outputs, the absence of experimental data may affect the reliability of the conclusions. Future research should validate the interaction networks experimentally to further confirm the co-degradation mechanisms between bones and muscles. Looking ahead, we plan to extend the study to MRI data analysis to further validate the role of CT values in disease progression while exploring the integrated application of other imaging data and biomarkers.[44] Additionally, longitudinal studies will help elucidate the dynamic changes in bone and muscle characteristics, providing more precise guidance for personalized treatment and early intervention.[45]

In conclusion, this study develops classification models to analyze bone-muscle interactions in osteoporosis, revealing synergistic degradation patterns of bone and muscle across disease stages. By introducing the innovative BMINet toolkit, we provide an efficient and interpretable framework for personalized analysis, offering a robust foundation to advance precision medicine and promote integrated care for osteoporosis patients.

## Data Availability

BMINet package is available on the Python Package Index (PyPI) platform, allowing users to easily download using the following command: pip install BMINet. The specific project page for the Python package can be found at https://pypi.org/project/bminet. Additionally, the source code and original data related to this study can be accessed for free at https://github.com/Spencer-JRWang/BMINet.

## Code Availability

*BMINet* package is available on the Python Package Index (PyPI) platform, allowing users to easily download using the following command: pip install BMINet. The specific project page for the Python package can be found at https://pypi.org/project/bminet. Additionally, the source code and original data related to this study can be accessed for free at https://github.com/Spencer-JRWang/BMINet.

## Author Contributions

J.W. (Jingran Wang) conceptualized the study, developed the machine learning models, and wrote the initial draft of the manuscript. Z.H. (Ziyan Hao), L.L. (Liyun Lin), and J.L. (Jiachen Liu) were responsible for data measurement and data curation. J.W. (Jiarong Wang) collected and organized the patient data. Z.T. (Zaixiang Tang) conducted the statistical analysis. D.G. (Dechun Geng) analyzed the application of the models and contributed to the interpretation of the results. C.N. (Caifang Ni) provided guidance on model development and methodological framework. H.Y. (Huilin Yang) contributed to the study design, mechanism analysis, and critical revision of the manuscript. K.L. (Kun Li) and J.D. (Jun Du) supervised the patient imaging examinations, ensured data quality control, provided guidance on modeling, and critically revised the manuscript. J.D., K.L. and H.Y. were responsible for verifying the underlying data reported in the manuscript. All authors reviewed and approved the final version of the manuscript and agree to be accountable for all aspects of the work.

## Compteting interests

The authors declare that they have no competing interests.

## Acknowledgements

This work was supported by the National Natural Science Foundation of China (Grant No. 82102609, 81701649, 82373688 and 81773541), Suzhou Basic Research Pilot Project (SSD2024084) and The First Affiliated Hospital of Soochow University Boxi Youth Natural Science Foundation (BXQN2023011). Funding was provided by K.L. (Grant No. 82102609) and J.D. (Grant Nos. 81701649, SSD2024084, and BXQN2023011).

---

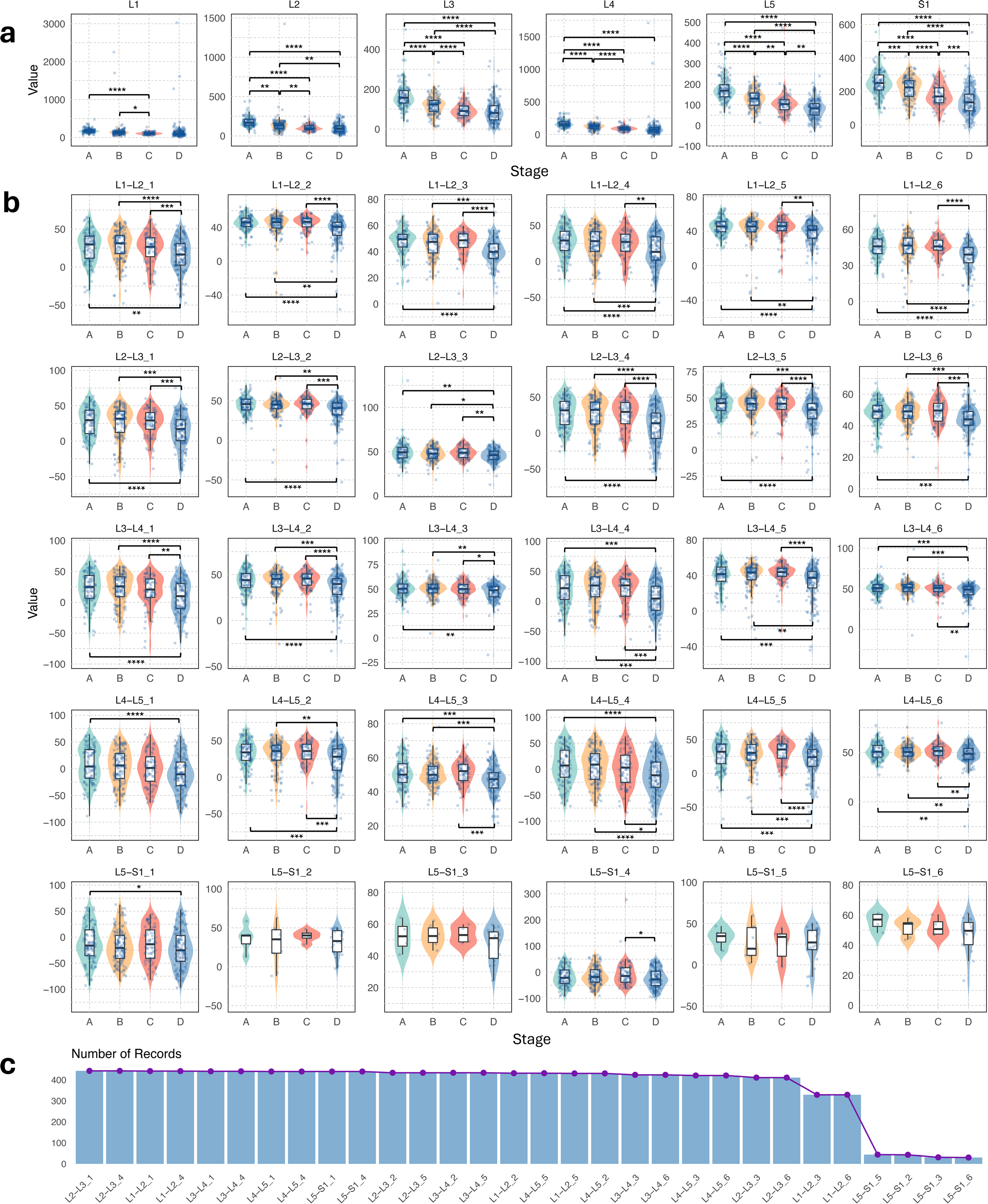

