## Supplementary Table 3 for "CT-based Osteoporosis Classification and Bone-Muscle Interaction Mapping Using Multiple Interpretable Machine Learning Models with the *BMINet* Framework"

| **Features** | **A vs B** | **A vs C** | **A vs D** | **B vs C** | **B vs D** | **C vs D** |
| --- | --- | --- | --- | --- | --- | --- |
| **L1** | 0.715 | 0.859 | 0.745 | 0.659 | *Not Sig* | *Not Sig* |
| **L2** | 0.714 | 0.856 | 0.829 | 0.658 | 0.669 | *Not Sig* |
| **L3** | 0.739 | 0.869 | 0.847 | **0.683** | 0.696 | *Not Sig* |
| **L4** | **0.749** | **0.876** | **0.877** | **0.688** | 0.739 | 0.618 |
| **L5** | **0.742** | **0.873** | **0.921** | 0.679 | **0.783** | 0.653 |
| **S1** | 0.614 | 0.752 | 0.849 | 0.674 | **0.774** | 0.650 |
| **L1-L2_2** | *Not Sig* | *Not Sig* | 0.690 | *Not Sig* | 0.6609 | 0.689 |
| **L1-L2_3** | *Not Sig* | *Not Sig* | 0.723 | *Not Sig* | 0.6662 | **0.714** |
| **L1-L2_6** | *Not Sig* | *Not Sig* | 0.713 | *Not Sig* | 0.7098 | **0.743** |
| **L2-L3_4** | *Not Sig* | *Not Sig* | 0.694 | *Not Sig* | 0.6757 | 0.675 |
| **L3-L4_5** | *Not Sig* | *Not Sig* | 0.632 | *Not Sig* | 0.6392 | 0.684 |

**Supplementary Table 3** **| Performance of key vertebrae and muscles across different groups.** The table records the top five features ranked by AUC in each group, along with their performance in other groups. Features with an AUC < 0.6 are marked as "Not Significant". The feature ranked first in each group is highlighted in red, while the feature ranked second is highlighted in blue.
