## Supplementary Table 4 for "CT-based Osteoporosis Classification and Bone-Muscle Interaction Mapping Using Multiple Interpretable Machine Learning Models with the *BMINet* Framework"

| **RFE Method** | **Features** | **CV Score** | **CV AUC**  **(95%CI)** | **Precision** | **Recall** | **F1** | **Time Cost** |
| --- | --- | --- | --- | --- | --- | --- | --- |
| *A vs B* |  |  |  |  |  |  |  |
| XGBoost | 12 | 0.670 | 0.703  (0.552 - 0.803) | 0.705 | 0.667 | 0.685 | 3min53s |
| LightGBM | 10 | 0.751 | 0.787  (0.719 - 0.844) | 0.759 | 0.806 | 0.782 | 1min58s |
| *A vs C* |  |  |  |  |  |  |  |
| XGBoost | 31 | 0.825 | 0.864  (0.814 - 0.896) | 0.765 | 0.785 | 0.775 | 2min26s |
| LightGBM | 8 | 0.858 | 0.881  (0.736 - 0.919) | 0.773 | 0.734 | 0.753 | 1min22s |
| *A vs D* |  |  |  |  |  |  |  |
| XGBoost | 26 | 0.851 | 0.905  (0.854 - 0.958) | 0.859 | 0.879 | 0.869 | 2min44s |
| LightGBM | 14 | 0.890 | 0.940  (0.905 - 0.976) | 0.887 | 0.894 | 0.891 | 1min36s |
| *B vs C* |  |  |  |  |  |  |  |
| XGBoost | 6 | 0.683 | 0.693  (0.495 - 0.788) | 0.527 | 0.494 | 0.51 | 3min29s |
| LightGBM | 13 | 0.697 | 0.701  (0.584 - 0.784) | 0.538 | 0.532 | 0.535 | 1min50s |
| *B vs D* |  |  |  |  |  |  |  |
| XGBoost | 15 | 0.698 | 0.783  (0.717 - 0.841) | 0.744 | 0.750 | 0.747 | 4min7s |
| LightGBM | 13 | 0.728 | 0.787  (0.678 - 0.894) | 0.734 | 0.689 | 0.711 | 2min9s |
| *C vs D* |  |  |  |  |  |  |  |
| XGBoost | 20 | 0.715 | 0.720  (0.644 - 0.822) | 0.708 | 0.773 | 0.739 | 3min21s |
| LightGBM | 19 | 0.716 | 0.752  (0.741 - 0.760) | 0.76 | 0.780 | 0.774 | 1min48s |

**Supplementary Table 4 | Comparison of XGBoost and LightGBM in RFE.**
